# Association between Healthcare Resources Inputs and Intravenous Tissue Plasminogen Activator Adherence Rate among Patients with Acute Ischemic Stroke

**DOI:** 10.1101/2023.05.25.23290558

**Authors:** Bo Kang, Suxi Zheng, Xin Yang, Chun-Juan Wang, Hong-Qiu Gu, Zi-Xiao Li, Yong-Jun Wang

## Abstract

**Background:** Intravenous Tissue Plasminogen Activator (IV rt-PA) significantly improves AIS patients’ functional outcomes within the treatment window, yet the usage of IV rt-PA among AIS patients are substantially lower in China than in developed countries. Healthcare resource utilization manages effective treatment patterns for patients who are adherent to IV rt-PA. This study investigates the association between healthcare resource inputs and IV rt-PA adherence and the impact of Gross Regional Product (GRP) on IV rt-PA.

**Methods:** 1,456 hospitals from 31 provinces with 158,003 acute ischemic stroke patients who had received IV rt-PA between 2015-2019 were recruited by the Chinese Stroke Center Alliance. The study outcome was the adherence rate of IV rt-PA in each hospital. Healthcare resource input was identified from three aspects: human, material, and economic. Multivariable linear regression was conducted by adjusting healthcare system characteristics and by further adjustment of GRP.

**Results:** The median (interquartile range) of IV rt-PA rate was 19.1% (8.6% -34.6%). Physician-nurse ratio (ß=0.023, p<0.001), nurse-bed ratio (ß=0.0343, p<0.001), and total health expenditure (ß=0.00002, p<0.001) were positively associated with the IV rt-PA adherence rate after controlling healthcare system factors. Through additional adjusting of GRP, only health expenditure was significantly positively associated with IV rt-PA adherence rate (ß=0.000018, p<0.001).

**Conclusions:** More health spending and being equipped with equally proportional physician-nurses and nurse-bed combinations in the provincial hospital will increase adherence to IV rt-PA among AIS patients. The difference in GRP among provinces may stimulate hospitals to provide more healthcare input from the workforce, thus indirectly increasing the usage of IV rt-PA.

## INTRODUCTION

Stroke is the leading cause of death and adult disability in China (1.7 million death with 38.6 million quality-adjusted life years (QALYs) in 2016)^1^. Intravenous tissue plasminogen activator (IV rt-PA) is one of the guideline-recommended treatments for patients with acute ischemic stroke (AIS) to improve the patient’s functional outcome^2^. However, adherence to IV rt-PA is low worldwide. Only 3% to 8.5% of potentially eligible patients receive IV rt-PA^3^. The adherence rate was especially unsatisfactory in China. Combined with statistical results from the China National Stroke Registry (CNSR,2007-2008) and China Quality Evaluation of Stroke Care and Treatment (China QUEST, 2007-2008), only 1.2–1.9% of AIS patients are treated with IV rt-PA^4^. This percentage is substantially lower than the adherence rate in the United States, Canada (8.1%-10.2%), and the Netherlands (6.4–14.6%)^5^.

Previous literature from several perspectives explained the low IV rt-PA rate in China. Administration deficiency factors included prehospital delay, the short time window in IV rt-PA, limited access to imaging examination, and a regional stroke care network shortage^6,7^. Economic burden factors comprised skyrocketing IV rt-PA cost and low insurance coverage^2, 8^. Current studies also illustrated that healthcare system factors were associated with a low compliance rate to IV rt-PA ^6,9,10,11^. It depends on physician skills and expertise (treatment by the neurologist, having a comprehensive stroke center) in the stroke division, the hospital’s staffing capacity (number of neurologists, number of having stroke team), and other organizational factors such as having coordination with emergency care and use of stroke-specific protocols.

Despite the discussion about the cause of low compliance for IV rt-PA, only some illustrated the contribution from the healthcare resource input field. Two studies investigated healthcare utilization and changes in functional status among stroke patients ^12, 13^. Emily O’Brien *et al*^14^ used healthcare-resourced data to examine the relationship between economic resources and In-hospital quality outcomes among AIS patients. However, no significant result was found. The uncertainty about how healthcare resources affect the adherence rate in IV rt-PA is still unknown. Further evidence of causal inference between IV rt-PA adherence rate and healthcare inputs from the workforce, facilities capacity, and economic scale is needed.

This study aims to bridge the knowledge gap and explore the variation of healthcare resource input and hospital system factors influencing nationwide quality improvement on stroke across China. Using data from the Chinese Stroke Center Alliance (CSCA), we sought to assess how healthcare resource input and hospital system characteristics influence the adherence rate to IV rt-PA in patients who arrived within 3.5 hours after symptom onset and were treated within 4.5 hours.

## METHODS

### Data Extraction

Hospital-level data were collected by the Chinese Stroke Association (CSA) from the CSCA database, a national, hospital-based, multi-center, voluntary, continuous quality assessment and improvement platform. The data coordinating center of the CSCA resides at the China National Clinical Research Center for Neurological Diseases (NCRC-ND), Beijing Tiantan Hospital. Provincial-level healthcare resource inputs were collected from the Yearbook of Health in the People’s Republic of China, National Bureau of Statistics of China from 2015-2019. Participating hospitals received a healthcare quality assessment and research approval to collect data in the CSCA without requiring individual patient informed consent under the common rule or a waiver of authorization and exemption from their Institutional Review Board.

### Study Population

The study included 1,456 hospitals that enrolled patients over 18 with AIS from secondary or tertiary hospitals in the CSCA across 31 provinces, autonomous regions, and municipalities in mainland China (excluding Hongkong, Taiwan, and Macau). Patients were recruited consecutively in all hospitals from Aug 2015 to July 2019. In addition, patients with medical contraindications (e.g., treatment intolerance, excessive risk of adverse reaction, patient/family refusal, or terminal illness/comfort care only) to intravenous thrombolysis in the time window were further excluded from the study. The final study population in this study was 1,456 hospitals with 158,003 AIS patients.

### Outcome Measure

The outcome measure was the adherence rate of IV rt-PA in each hospital. The adherence rate was calculated using the number of AIS patients receiving IV rt-PA within 4.5 hours divided by the number of AIS patients who were eligible for IV rt-PA and were without any medication contradictions, including treatment intolerance, excessive risk of adverse reaction, patient/family refusal or terminal illness/comfort care only.

### Multilevel

This study included provincial-level healthcare resource inputs, provincial-level hospital gross regional product per capita, and hospital-level healthcare system characteristics.

### Healthcare Resources Inputs

We categorized provincial-level health resource inputs into human, material, and economic aspects(Table 1). For human aspects, we measured the ratio of physician to nurse; the ratio of nurse to bed; the number of physicians per 1,000 population; and the number of nurses per 1,000 population. The physician-nurse ratio was directly measured by the number of board-certified physicians divided by the number of registered nurses (RNs) in each province. Similarly, we measured the nurse-bed ratio by dividing the number of RNs by the number of medical beds in each province. We calculated the number of physicians per 1,000 by dividing the number of physicians by the total population in each province and multiplying by 1,000.

**Table 1.** Provincial-level healthcare resource inputs and GRP measurement table.

| Healthcare resource inputs | Algorithm |
| --- | --- |
| Physician-nurse ratio | $\frac{\# \text{ of Physicians}}{\# \text{ of RNs}}$ |
| Nurse-bed ratio | $\frac{\# \text{ of RNs}}{\# \text{ of Medical beds}}$ |
| Physician numbers, per 1,000 | $\frac{\# \text{ of Physicians}}{\text{Total Population}} \times 1000$ |
| Nurse numbers, per 1,000 | $\frac{\# \text{ of RNs}}{\text{Total Population}} \times 1000$ |
| Medical institute numbers, per 1,000 | $\frac{\# \text{ of Medical institutes}}{\text{Total Population}} \times 1000$ |
| GRP | $\frac{\text{GDP}}{\text{Total Population}}$ |

Source: Yearbook of Health in the People's Republic of China, published by the National Bureau of Statistics of China, 2015-2019<sup>15-19</sup>.
All healthcare resource inputs and GRP were measured yearly at the provincial level.
The total population and GDP were calculated in each province.

Similarly, the index for the number of RNs per 1,000 population was calculated by dividing the sum of the number of RNs in each province by the province population and multiplying by 1,000. We employed the same algorithm to measure medical institutes per 1,000 population as representative of material-related input. Economic-related input was measured by each province’s total expenditure on healthcare services.

All data used to quantify healthcare resource inputs were collected from the Yearbook of Health in the People’s Republic of China, published by the National Bureau of Statistics of China,2015-2019^15-19^.

### Healthcare System Characteristics/Covariates

We used the provincial-level hospital’s gross regional product per capita (GRP), collected from the National Bureau of Statistics of China, to represent the macroeconomic index of the healthcare system in each province. The GRP was measured by dividing the GDP in each province by the province’s population. As the existing body of literature and human development report described, GRP is conceptually equivalent to gross domestic product but was measured at the hospital’s province level. Other healthcare system characteristics were simplified into three categories: hospital training skill and expertise, hospital facility and staffing, and organizational elements^11^. We listed variables of hospital system factors based on experience from previously published articles ^11, 20, 21^, but the final candidate variables rely on both literature support and model fitting performance. We identified a hospital training skill and expertise by hospital level (1 if the hospital is a tertiary hospital and 0 if the hospital is a secondary hospital); the presence of stroke certification (1 if the hospital is a comprehensive stroke center (CSC);0 if the hospital is a primary stroke center (PSC))^11^. Hospital facility and staffing were proxied by the number of beds in the neurology department (NEU), the number of neurologists, and whether a hospital has an emergency department (E.D.). We included whether the hospital cooperates with 120 emergency center (120-EC.) and whether the hospital provides E.D. services for neurological treatment as proxies for organizational factors. All variables had a low proportion of missing data (<3%) due to missing completely at random (MCAR), which will not affect the validity of the study results.

### Statistical Analysis

Adherence rates were initially calculated in quartiles at the hospital level and then aggregated at the province level to determine what human resource input might be associated with adherence rate to IV rt-PA. Summary statistics of hospital characteristics were computed within each adherence rate quartile.

Multivariable linear regression was conducted to identify the effect of each factor. All factors were included in the same model, and a stepwise (the significant level of 0.05 for removal from the model) variable selection methodology was implemented in multilevel model analysis. The results of regression were presented as the crude beta coefficient, beta coefficient adjusted by the healthcare system factors (Stroke Centre type, Hospital Grade, Neurologist number, E.D with acute ischemic stroke team, Presence of E.D, cooperating with E.D, Hospital bed size in NEU, and year), and beta coefficient adjusted by above covariates plus GRP with 95% confidence interval(95-CI). Beta coefficients described the magnitude and change in the adherence rate to IV-rt PA for every one-unit change in the specific healthcare resource input while holding other factors constant. The standardized(adjusted) beta coefficients were used to compare the effect of each element on the adherence rate after scaling the different units of factors. P<0.05 was used as the rule of thumb for the significance level. All statistical analyses were performed with the R software (V3.6.3).

## RESULTS

The adherence rate of IV rt-PA of a total of 1,456 unique hospitals with 158,003 patients from 31 provinces in Mainland China, except for Hong Kong Special Administrative Region (HKSAR), Macau SAR, and Taiwan between 2015-2019, were studied along with quartile level.

### Baseline Characteristics by IV rt-PA with Each Quartile

Healthcare resource inputs and certain hospital characteristics among the adherence rate quartiles are illustrated in Table 2. The IV rt-PA rate at each quartile was 8.6%(25th), 19.1%(50th), 34.6%(75th), and 34.6% above, respectively. Table 1 results illustrated that hospitals with high adherence rates primarily presented high input on human, material, and economic resources. The trend of medical expenditures among the quartiles kept increasing. The value of total expenditure among hospitals was ¥1,968 billion in the first quartile(Q1), compared to ¥2,300 in the second quartile, ¥2,495 in the third quartile, and reach to ¥2,680 in the fourth quartile. The mean value of total expenditures on health per hospital was 2,300 hundred million for AIS patients. Likewise, hospitals with high adherence rates also performed high physician-nurse and nurse-bed ratios, while the difference was not large. The Mean value of the physician-nurse and nurse-bed ratios was 0.86 and 0.48. The highest quartile level (99th) of physician-nurse and the nurse-bed rate was close to 1:1 and 1:2. Physician number rate per 1,000 population varies by quartiles, but it all around at the 2 with significant difference(P<0.001). Medical institute numbers with the lowest mean volume per 1,000 population (0.66) provided unexpectedly high performance in the fourth quartile(Q4), compared with 0.71 in Q3, 0.78 in Q2, and 0.78 in Q1.

**Table 2.** Baseline hospital characteristics overall by IV rt-PA quartile.

|  | IV rt-PA quartile by hospital number, the median adherence rate |  |  |  |  | P-value * |
| --- | --- | --- | --- | --- | --- | --- |
|  | Overall<br>(n=1,456) | Quartile<br>1<br>< 8.6%<br>(n=364) | Quartile2<br>8.6%-<br>19.1%<br>(n=364) | Quartile3<br>19.1%-<br>34.6%<br>(n=364) | Quartile4<br>>34.6%<br>(n=364) |  |
| **Physician-nurse ratio** | 0.86<br>(0.32) | 0.80<br>(0.21) | 0.83<br>(0.28) | 0.92<br>(0.39) | 0.91<br>(0.37) | <0.001 |
| **Nurse-bed ratio** | 0.48<br>(0.08) | 0.46<br>(0.07) | 0.46<br>(0.06) | 0.49<br>(0.08) | 0.50<br>(0.09) | <0.001 |
| **Physician numbers, per 1,000** | 2.07<br>(0.37) | 2.03<br>(0.34) | 2.00<br>(0.27) | 2.08<br>(0.39) | 2.16<br>(0.44) | <0.001 |
| **Nurse numbers, per 1,000** | 2.67<br>(0.42) | 2.66<br>(0.38) | 2.64<br>(0.33) | 2.67<br>(0.45) | 2.72<br>(0.49) | 0.139 |
| **Medical institute numbers, per1,000** | 0.73<br>(0.25) | 0.78<br>(0.24) | 0.76<br>(0.24) | 0.71<br>(0.25) | 0.66<br>(0.25) | <0.001 |
| **Total expenditure on health service** | 2356.29<br>(1105.12) | 1968.87<br>(969.09) | 2300.46<br>(1053.22) | 2494.86<br>(1175.11) | 2680.35<br>(1096.14) | <0.001 |
| **GRP per capita, mean (SD), ¥** | 5.89<br>(2.23) | 5.16<br>(1.78) | 5.48<br>(1.95) | 6.20<br>(2.18) | 6.80<br>(2.60) | <0.001 |
| **Tertiary hospital (n, %)** | 583<br>(59.5%) | 118<br>(48.2%) | 149<br>(58.0%) | 163<br>(66.3%) | 153<br>(65.9%) | <0.001 |
| **Comprehensive stroke Centre (n, %)** | 223<br>(22.8%) | 32<br>(13.1%) | 51<br>(19.8%) | 70<br>(28.5%) | 70<br>(30.2%) | <0.001 |
| **Hospital bed-size in NEU** | 88 | 85 | 88 | 92 | 88 | 0.653 |
| **Neurologists, mean** | 15 | 13 | 15 | 17 | 17 | 0.001 |
| **Hospital with E.D (n, %)** | 967<br>(99.1%) | 238<br>(97.5) | 257<br>(100.0) | 242<br>(98.8) | 230<br>(100.0) | 0.011 |
| **Cooperation with 120-EC (n, %)** | 938<br>(96.2%) | 228<br>(93.8%) | 254<br>(98.8%) | 234<br>(95.5%) | 222<br>(96.5%) | 0.029 |
| **Hospital with E.D service for neurological treatment (n, %)** | 948<br>(98.3%) | 234<br>(96.7%) | 252<br>(98.8%) | 238<br>(98.8%) | 224<br>(99.1%) | 0.141 |

P-values are computed using the Cochran-Armitage test for categorical variables and the Jonckheere-Terpstra trend test for continuous variables.
The adherence rate to IV rt-PA was estimated using the number of AIS patients receiving IV rt-PA divided by the number of AIS patients eligible for IV rt-PA.
GRP, gross regional product; E.D., emergency department; NEU, neurology department

Significant differences were also identified in healthcare system factors. Like the trend of total expenditure on health, the macroeconomic index GRP per capita kept increasing from 5.16 in Q1 to 6.80 in Q4. For elements reflecting hospital training, skills, and expertise in AIS treatment, the number of tertiary hospitals varied among the quartiles, but its percentage seemed kept increased from 48.2% (Q1) to 65.9%(Q4). Hospital bed size in NEU was an average of 90 units at each hospital treated with AIS patients. For the characteristics reflecting facility and staffing, Hospitals with higher adherence rates of IV rt-PA had more neurologists and a higher percentage of stroke centers than those with lower IV rt-PA rates. In addition, the IV rt-PA compliance rate across the quartile varied in the portion of the hospital with E.D. (p=0.011) and hospitals with the cooperation of 120-EC(p=0.021), and in E.D. service of neurologist service(p=0.141). Such insignificant differences may imply that organizational factors did not directly influence the adherence rate to IV rt-PA.

### Multivariable Analysis of Adherence Rate to IV rt-PA and Factors

The results of multivariable regression are reported in Table 3. We take Hospital-level characteristics, including GRP per capita, stroke center type, hospital Grade, neurologist number,

**Table 3.** Multivariable regression analysis between IV rt-PA and influential factors.

| **Parameter** | **Crude**<br>**Estimate(β) (95% CI)** | **Adjusted\***<br>**Estimate(β) (95% CI)** | **Adjusted\*\***<br>**Estimate(β) (95%**<br>**CI)** |
| --- | --- | --- | --- |
| **Physician-nurse ratio** | 0.026 (0.016-0.035) | 0.023 (0.009-0.037) | 0.003 (-0.012-0.018) |
| **Nurse-bed ratio** | 0.253 (0.181-0.326) | 0.343 (0.239-0.447) | 0.070 (-0.063-0.202) |
| **Medical institute numbers, per1,000** | -0.146 (-0.177- -0.115) | -0.030 (-0.067- 0.007) | 0.021 (-0.019-0.061) |
| **Total expenditure on health service** | 0.00004 (0.00003-0.00005) | 0.00002 (0.000016-0.007) | 0.000018 (0.000010-0.000025) |

\*Adjusted hospital-level characteristics (Stroke Centre type, Hospital Grade, Neurologist number, emergency departments with acute ischemic stroke team, presence of E.D, Cooperating with E.D, hospital bed size in NEU, and year)
\*\*Adjusted hospital-level characteristics and GDP per capita、

E.D with acute ischemic stroke team, presence of E.D, cooperating with 120-E.C, Hospital bed size in NEU, and year (2015 as reference year) in the model as a confounding variable. Multivariable regression results were described in three models: the model with crude beta estimation, the adjusted model with controlling hospital characteristics, and the adjusted model with controlling for hospital characteristics and GRP. Four healthcare resource inputs, physician-nurse ratio, nurse-bed ratio, medical institute number, and total expenditure on healthcare remain significantly associated with adherence rate to IV rt-PA. The crude beta coefficient for four healthcare resource inputs were 0.026 in the physician-nurse ratio,0.253 in the nurse-bed ratio, - 0.146 in medical institutes per 1000 population, and 0.00004 in total expenditure on health service. After controlling hospital-level characteristics (without GRP per capita), the Adherence rate increases by 3.4% with a 1% increase in nurse-bed ratio and 2.3% with a 1% increase in physician-nurse ratio.

Moreover, every extra 100 million Chinese yuan investment in health expenditure was associated adherence rate increase of 0.002%. Medical institute number per 1,000 population was no longer associated with adherence rate on IV rt-PA. After further adjustment of GRP per capita and other healthcare system characteristics, the only significant factor associated with adherence rate to IV rt-PA was total expenditure on health service. The adherence rate of provincial hospitals with an extra 100 million investment in health services was 0.0018% higher than those without spending additional cost on medical services.

## DISCUSSION

This national, multi-level study not only observed significant differences in adherence rates among 1,456 hospitals in 31 provinces from Mainland China but also illustrated that human and economic resources positively affected adherence to IV rt-PA, even after controlling for healthcare system characteristics. Medical institute numbers per 1,000 population gradually decreased with the adherence rate to IV rt-PA increased. One explanation is that provinces with higher adherence rates usually contribute larger GRP but also afford more population than those with lower adherence rates. Medical institute numbers per 1,000 were high depending on the total population of each province but were not associated with IV rt-PA adherence rate. Our multivariate analysis also confirmed this interpretation. Total expenditure on health becomes the only contributor to the disparity of the adherence rate after adjusting the GRP difference in provinces. This evidence suggests that increasing healthcare expenditure improves hospital adherence rates to IV rt-PA.

After further control of GRP, the IV rt-PA rate did not significantly influence human and material resources. This result indicated that the input level of those resources was heavily dependent on the development of GRP per capita or vice versa. Hospitals in provinces with larger GRP per capita would recruit more skilled physicians and nurses for the growing population and establish more medical organizations than those in lower GRP provinces^22^.

The correlation analysis between healthcare resources and GRP further supports this argument (Table 4). The correlation coefficient between GRP per capita and nurse-bed ratio, physician-nurse-ratio, medical institutes number per 1,000 population, and total health expenditure are 0.54, -0.51, 0.15, and 0.45, respectively. These numbers indicate that GRP positively correlated with the nurse-bed ratio and negatively correlated with the medical institute’s number. The existing literature also provides strong evidence. GDP per capita growth can occur through productivity changes, saving, investment(economic), or labor supply (human and material capital). One example is healthier workers (i.e., physicians or nurses), usually expected to use their working time and resources better, directly increasing productivity^23^. Human and material capital gains will raise productivity and per capita GDP ^24^. All the previous efforts and our findings reveal the cointegrating relationship between GRP per capital and healthcare resources input.

**Table 4.**
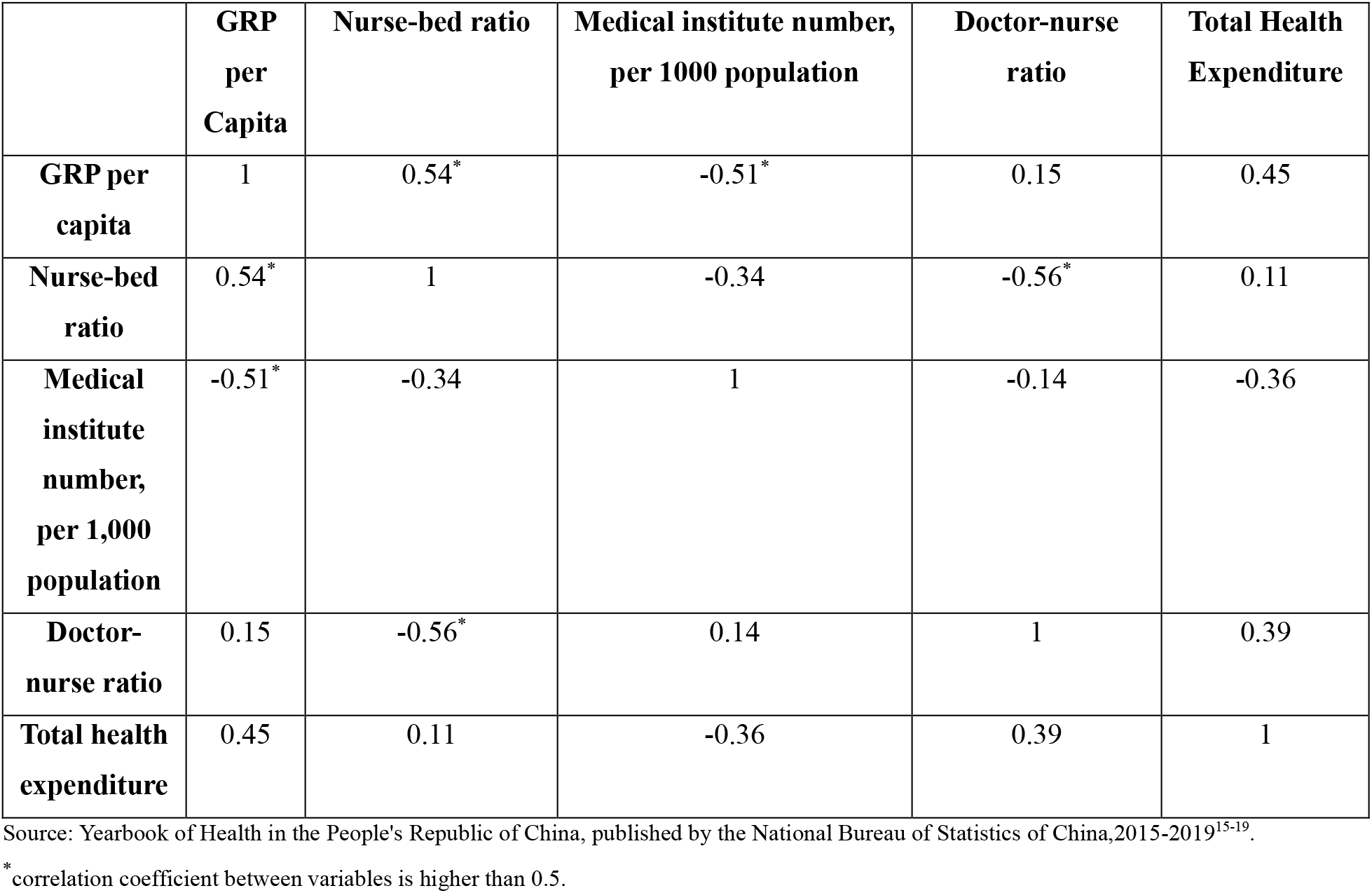
Provincial-level healthcare resource inputs correlation matrix.

|  | **GRP per Capita** | **Nurse-bed ratio** | **Medical institute number, per 1000 population** | **Doctor-nurse ratio** | **Total Health Expenditure** |
| --- | --- | --- | --- | --- | --- |
| **GRP per capita** | 1 | 0.54* | -0.51* | 0.15 | 0.45 |
| **Nurse-bed ratio** | 0.54* | 1 | -0.34 | -0.56* | 0.11 |
| **Medical institute number, per 1,000 population** | -0.51* | -0.34 | 1 | -0.14 | -0.36 |
| **Doctor-nurse ratio** | 0.15 | -0.56* | 0.14 | 1 | 0.39 |
| **Total health expenditure** | 0.45 | 0.11 | -0.36 | 0.39 | 1 |

Source: Yearbook of Health in the People's Republic of China, published by the National Bureau of Statistics of China,2015-2019<sup>15-19</sup>.
\* correlation coefficient between variables is higher than 0.5.

The statistics show that the IV rt-PA treatment adherence rate will increase by 0.002% if the Chinese government’s additional cost is 100 million CNY (Chinese yuan). While a significant investment may lead to only a minimal improvement in the short term, increasing total health expenditure may significantly impact IV rt-PA use in the long run. Early evidence indicates that IV rt-PA treatment within 4.5 hours is more cost-effective for AIS patients in the long run than in the short run. The estimate of 38 Organization for Economic Cooperation and Development (OECD) countries described that using IV rt-PA within 4.5 hours could cause a 0.101 gain of Quality Adjusted Life Years (QALYs) with an additional CNY 9520(1,460USD) in two years. However, it will lead to a long-term increase of 0.422 QALYs at an additional CNY6,530 (about 1,000 USD) in thirty years ^8^. Economic resource input (health expenditure on GRP) is still some years from reaching its maximum effect. The impact of expanding the IV rt-PA rate is better executed in the long period than in a couple of years. Health improvement results in long-term sustainable growth in GDP. In most OECD countries, one increase in life expectancy will lead to gains in productivity and result in a 5% increase in GDP per capita ^24^. The increased GDP, in turn, will keep expanding patients’ life-saving years by transiting more proportion of GDP on health expenditure. Meanwhile, as hospitals in economically developed provinces have more capability of acquiring financial support and reimbursement from government funding than those in underdeveloped regions, they are inclined to be equipped with more experienced neurologists, more comprehensive emergency services, emergency services routine coordination with local emergency centers performed better in adhering to the delivery of IV rt-PA^11^. In this positive cycle of stroke care, increasing GRP per capita will stimulate government spending on additional healthcare costs to expand IV rt-PA use, thus achieving optimal health outcomes.

The impact of emergency medical services and ED on the thrombolysis rate is vital. While Acar et al.^20^ and Zheng et al.^11^ found that equipped with medical services and an organized emergency department would greatly facilitate performance in the delivery of IV rt-PA treatment^11, 20^, thus increasing IV rt-PA use, J.D.H. Van Wijngaarden et al. ^25^ found that triage emergency rooms and ambulance services show no significant association with thrombolysis rates^25^. Our study further supports such conclusion and evidence that there is no clear association (negative or positive) between IV rt-PA use and organization factors with E.D. use. Such controversy may be explained by the fear of emergency department physicians about using IV rt-PA. Since most stroke patients are first treated by emergency medical staff, E.D. physicians’ decision to administer IV rt-PA to patients with ischemic stroke is crucial. A national survey found that 40% of emergency physicians would not use IV rt-PA due to the risk of intracerebral hemorrhage, clinical ambiguity, and concern about expensive cost ^26^. Such fear and risk of using IV rt-PA can be mitigated. Increased hospital reimbursement may make new training programs possible, which will help these emergency physicians feel as confident about treating patients with stroke as they are about treating patients with myocardial infarction.

There are several limitations to our study. First, a large group of participants from CSCA was from the secondary and tertiary hospitals instead of the primary hospital. Hospitals with higher classes usually recruit more physicians and nurses and acquire larger healthcare budgets than those with lower classes. Second, although healthcare system characteristics were comprehensively controlled in this study, some unmeasured, unobserved factors not considered might also affect the IV rt-PA adherence rate. Patients may be reluctant or refuse to receive IV rt-PA treatment due to fear of existing side effects, including a short time window, low recanalization, and high risk of intracranial hemorrhage. However, patients’ tendencies could not be measured due to data limitations. Third, IV rt-PA is one of the most critical QI indices authorized by Get With the Guideline for stroke care. Still, it neither replaces the role of the other ten indexes in stroke treatment nor has outstanding performance for the treatment of hemorrhagic stroke patients, thus limiting the study’s generalizability to AIS patients only.

Our finding provided evidence that the low adherence rate in China may be attributed to insufficient input on human-resource health expenditure. Health policymakers and hospital managers should pay attention to increasing funding for health expenditures. Meanwhile, the government should also provide a specific preferential policy that encourages recruiting more well-trained, certified physicians and nurse practitioners to compensate for the insufficiency of human resources in the current medical workforce. All these implementations are beneficial to expand IV rt-PA compliance among eligible AIS patients.

## CONCLUSIONS

The adherence rate of IV rt-PA increased with the doctor-nurse ratio, nurse-bed ratio, and total expenditure on health without adjustment of GRP. Stimulating and integrating the relationship between GRP per capita and human and material resource input is critical to improving IV rt-PA adherence rate, especially considering medical institutes’ capacity and expertise disparity in the under-developed provinces.

## Data Availability

Hospital-level data were collected by the Chinese Stroke Association (CSA) from the CSCA database, a national, hospital-based, multi-center, voluntary, continuous quality assessment and improvement platform. The data coordinating center of the CSCA resides at the China National Clinical Research Center for Neurological Diseases (NCRC-ND), Beijing Tiantan Hospital. Data are available upon reasonable request. The datasets used and/or analysed during the current study are available from the corresponding author on reasonable request.

